# Impact of SARS-CoV-2 vaccination of children ages 5-11 years on COVID-19 disease burden and resilience to new variants in the United States, November 2021-March 2022: a multi-model study

**DOI:** 10.1101/2022.03.08.22271905

**Authors:** Rebecca K. Borchering, Luke C. Mullany, Emily Howerton, Matteo Chinazzi, Claire P. Smith, Michelle Qin, Nicholas G. Reich, Lucie Contamin, John Levander, Jessica Kerr, J Espino, Harry Hochheiser, Kaitlin Lovett, Matt Kinsey, Kate Tallaksen, Shelby Wilson, Lauren Shin, Joseph C. Lemaitre, Juan Dent Hulse, Joshua Kaminsky, Elizabeth C. Lee, Jessica T. Davis, Kunpeng Mu, Xinyue Xiong, Ana Pastore y Piontti, Alessandro Vespignani, Ajitesh Srivastava, Przemyslaw Porebski, Srini Venkatramanan, Aniruddha Adiga, Bryan Lewis, Brian Klahn, Joseph Outten, Benjamin Hurt, Jiangzhuo Chen, Henning Mortveit, Amanda Wilson, Madhav Marathe, Stefan Hoops, Parantapa Bhattacharya, Dustin Machi, Shi Chen, Rajib Paul, Daniel Janies, Jean-Claude Thill, Marta Galanti, Teresa Yamana, Sen Pei, Jeffrey Shaman, Guido Espana, Sean Cavany, Sean Moore, Alex Perkins, Jessica M. Healy, Rachel B. Slayton, Michael A. Johansson, Matthew Biggerstaff, Katriona Shea, Shaun A. Truelove, Michael C. Runge, Cécile Viboud, Justin Lessler

## Abstract

**Background:** SARS-CoV-2 vaccination of persons aged 12 years and older has reduced disease burden in the United States. The COVID-19 Scenario Modeling Hub convened multiple modeling teams in September 2021 to project the impact of expanding vaccine administration to children 5-11 years old on anticipated COVID-19 burden and resilience against variant strains.

**Methods:** Nine modeling teams contributed state- and national-level projections for weekly counts of cases, hospitalizations, and deaths in the United States for the period September 12, 2021 to March 12, 2022. Four scenarios covered all combinations of: 1) presence vs. absence of vaccination of children ages 5-11 years starting on November 1, 2021; and 2) continued dominance of the Delta variant vs. emergence of a hypothetical more transmissible variant on November 15, 2021. Individual team projections were combined using linear pooling. The effect of childhood vaccination on overall and age-specific outcomes was estimated by meta-analysis approaches.

**Findings:** Absent a new variant, COVID-19 cases, hospitalizations, and deaths among all ages were projected to decrease nationally through mid-March 2022. Under a set of specific assumptions, models projected that vaccination of children 5-11 years old was associated with reductions in all-age cumulative cases (7.2%, mean incidence ratio [IR] 0.928, 95% confidence interval [CI] 0.880-0.977), hospitalizations (8.7%, mean IR 0.913, 95% CI 0.834-0.992), and deaths (9.2%, mean IR 0.908, 95% CI 0.797-1.020) compared with scenarios where children were not vaccinated. This projected effect of vaccinating children 5-11 years old increased in the presence of a more transmissible variant, assuming no change in vaccine effectiveness by variant. Larger relative reductions in cumulative cases, hospitalizations, and deaths were observed for children than for the entire U.S. population. Substantial state-level variation was projected in epidemic trajectories, vaccine benefits, and variant impacts.

**Conclusions:** Results from this multi-model aggregation study suggest that, under a specific set of scenario assumptions, expanding vaccination to children 5-11 years old would provide measurable direct benefits to this age group and indirect benefits to the all-age U.S. population, including resilience to more transmissible variants.

## Introduction

SARS-CoV-2 vaccines have contributed to reducing serious outcomes of COVID-19, including severe disease, hospitalization, and death in the United States in 2021 [Scobie et al. MMWR]^1^. COVID-19 vaccination started in late December 2020, and demand largely surpassed supply through the early months of 2021^2^. Groups at higher risk, including health care workers and individuals aged ≥65 years, were prioritized to receive SARS-CoV-2 vaccines first. To further direct the first vaccine doses to those most at risk, on December 20, 2020, the Centers for Disease Control and Prevention (CDC), Advisory Committee on Immunization Practices [ACIP] recommended that COVID-19 vaccine initially be offered to persons aged ≥75 years and non– health care frontline essential workers^3^ <u>[Dooling et al. REF]</u>. Vaccine uptake, particularly in persons aged ≥75 years, increased quickly, contributing to a shift in the age distribution of severe cases by April 2021^4,5^ [<u>Boehmer et al. MMWR</u>, <u>Malmgren et al.</u>]. Vaccine emergency-use authorization was expanded by the U.S. Food and Drug Administration (FDA) to include persons 16 years of age or older in April 2021, and persons aged 12-15 years on May 10, 2021^6^ <u>[FDA approval site]</u>.

Despite encouraging signs of a receding pandemic in the U.S. in spring and early summer 2021, emergence of the Delta variant led to renewed COVID-19 risk overall, and particularly among children <12 years old, for whom the vaccine was not yet available. For example, COVID-19 hospitalizations per 100,000 in the U.S. increased among children and adolescents from 0.3 in June to 1.4 in late August 2021^7^ [Siegel et al.]; these surges were likely driven by increased transmissibility and severity of the Delta variant^8,9^ [Delahoy et al., Sonabend et al.]. Thus, despite relatively high coverage in the previously eligible population (approximately 65% of eligible individuals in the U.S. receiving two doses of primary series [Pfizer and Moderna] or one dose of Janssen vaccines by September 2021 <u>[REF]</u>^2^), there was reason to believe that the expansion of vaccination to children 5-11 years old could have an appreciable impact on the U.S. epidemic. However, the magnitude of this impact remained unclear given overall trends in acquired immunity, potential differences in age-specific transmission, and the size of this group (7.6% of the U.S. population).

In anticipation of authorization and recommendations for expanding vaccination to children 5-11 years old in the U.S. (e.g., FDA and CDC)^10^ <u>[REF]</u>, members of the COVID-19 Scenario Modeling Hub undertook a multiple-model approach to assess potential effects of immunizing children 5-11 years old (prior to authorization on October 29 2021^11^ <u>[REF]</u>). Our findings include ensemble projections, which aggregate over the individual models, of COVID-19 cases, hospitalizations, and deaths through March 2022.

## Methods

### Overview of Scenario Hub and epidemiological assumptions

The COVID-19 Scenario Modeling Hub (http://covid19scenariomodelinghub.org) was established in December 2020 to generate 6-month-ahead projections of the COVID-19 trajectory in the U.S. under scenarios capturing different epidemiological and intervention assumptions^12^ <u>[REF]</u>. For each round of scenarios, multiple modeling groups were convened in an open call to produce projections of weekly cases, hospitalizations, and deaths at both the state- and national-levels. The round discussed in this paper (the ninth round) focused on childhood vaccination, with team-specific projections made by nine modeling teams (see Supplement for complete list of team names) for the period September 12, 2021 to March 12, 2022. Round 9 considered four scenarios, including scenarios with and without vaccination in children 5-11 years old with administration beginning on November 1, 2021 in the U.S., and with and without the emergence of a hypothetical more transmissible variant in the U.S. on November 15, 2021^13^ <u>[REF github]</u>. The hypothetical variant was assumed to be 50% more transmissible than viruses circulating at the start of the projection period (see Table 1 for additional scenario details).

**Table 1:** Specification of the four scenarios included in the COVID-19 Scenario Modeling Hub’s Round 9 projections. The four scenarios (A, B, C, and D) differed in whether or not they included vaccination of children 5-11 years old, and whether or not a new, more transmissible variant emerged.

|  |  |  |
| --- | --- | --- |
|  | The same mix of variants circulate throughout the projection period.<br>No change in virus transmissibility. | A more transmissible variant emerges, comprising 1% of circulating viruses on **Nov 15**. The new variant is **1.5X** as transmissible as viruses circulating at the beginning of the projection period. |
| Vaccination among 5-11 year-olds is approved and immunization begins on Nov 1. Each state's uptake rate reflects the percent coverage increases observed for 12-17 year-olds since distribution began on May 13. | A | C |
| No vaccination for children under 12 years. | B | D |

In scenarios with vaccination of children 5-11 years old, uptake rates in children were specified to reflect those reported by the CDC for 12-17 year-olds at the state level since administration began for that age group on May 13, 2021^2^ <u>[REF]</u> (i.e., assumed to match the state-specific uptake curve in older children offset to begin November 1, 2021). Projected uptake in individuals over 12 years was at each team’s discretion, but informed by Pulse and CovidCast hesitancy surveys^14,15^. State-level vaccination coverage for older individuals was implemented to reflect reports from the CDC^2^. Age-group specific resolution depended on each team’s modeling assumptions, but saturation levels were assumed to be consistent with observed data. Booster vaccines were not included in order to focus on the potential effects of vaccination in children 5-11 years old.

Delta-variant specific vaccine effectiveness (VE) against infection, symptoms, and severe outcomes also were at team discretion for all age groups (see Supplemental Table 1); although estimates based on U.S. and U.K. studies were provided for guidance^16-19^ (<u>REACT</u> study, <u>Bernal et al.</u>, <u>Rosenberg et al MMWR</u>, <u>PHE surveillance report</u>). In the absence of VE estimates for children when scenarios were developed, VE in children 5-11 years old was assumed to match those of older age groups. Vaccine effectiveness estimates were also assumed to remain the same for the hypothetical new variant as for those assumed for the Delta variant. Additional assumptions were at the discretion of teams, based on their best scientific judgment, so that uncertainty in these areas would be reflected in the projections. This included assumptions about seasonal effects, including school terms, non-pharmaceutical interventions, and waning immunity. Details on these assumptions for each team are provided in meta-data available in the data repository associated with this study^13^ (also see Supplemental Table 2). Data through September 11, 2021 were used for model calibration.

### Ensemble estimates

Probabilistic projections (see Supplemental Methods) were reported by each of the nine modeling teams for each scenario, outcome metric, location, and week over the 26-week projection period (September 12, 2021-March 12, 2022). Ensemble projections were generated using a trimmed linear opinion pool method, where cumulative probabilities are averaged at a given value across all of the distributions submitted by the modeling groups, with the highest and lowest probabilities excluded before averaging^20,21^ [Stone 1961, Jose 2014]. Weekly ensemble distributions were used to create time series projections (Figure 1) for incident and cumulative outcomes.

**Figure 1.**
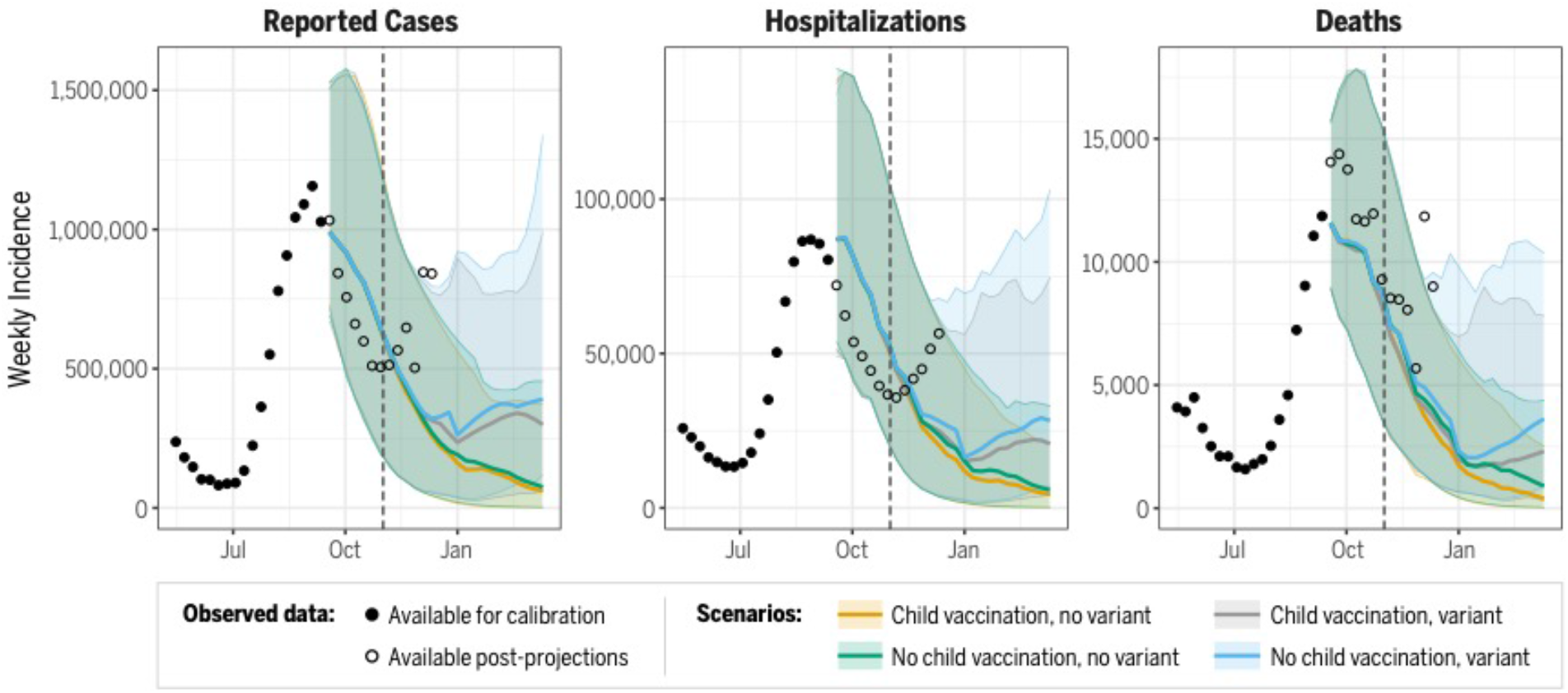
Projected cases, hospitalizations, and deaths for all ages at the national-level (September 12, 2021 – March 12, 2022). Median ensemble projections for each scenario displayed as lines, 95% projection intervals displayed as shaded regions. Observed ground truth data available for model calibration (solid points) and after model fitting (open circles) through December 11, 2021 (after which Omicron became prevalent in the US, departing from specified scenarios). The start date for vaccination of children 5-11 years old, November 1, 2021, is indicated by a dashed line.

### Evaluation of the benefits of vaccine expansion to children 5-11 years old

To summarize the projected overall benefits of the vaccine program expansion across all nine modeling teams, we used a standard meta-analytic approach with random effects [<u>REF</u>]^22^. Briefly, for each model and location (nationally and for all 50 states individually) we estimated the mean difference in cumulative incidence and the mean incidence ratio between scenarios with and without children 5-11 years old vaccinated, stratified by presence or absence of the new variant, for the portion of the projection period following the assumed start date of childhood vaccination (i.e., from November 1, 2021). Model-specific variance for the mean difference was estimated as the sum of the variances at each timepoint, while variance of the incidence ratio was estimated using the delta method^23^; both methods were scaled to standard errors using the number of replicates. Model-specific estimates and standard errors were combined via random effects meta-analysis using restricted maximum likelihood (REML); the same procedures were followed for best approximating the direct effect of vaccine expansion (within the younger age group that most closely matched the 5-11-year-old population, e.g., 0-11 year-olds, 5-11 year-olds, projections submitted by five teams). Full details of the methodological approach are found in the supplementary methods section.

All projections and code for reproducing results are publicly available at https://github.com/midas-network/covid19-scenario-modeling-hub.

## Results

### Overall trajectory

The ensemble projected a declining COVID-19 incidence from September through December 2021 (Figure 1). While national projections from different models largely agreed on trends and aligned with a decline in national case data through October 2021, substantial quantitative uncertainty remained (Figure S1). The median national projections did not follow the trend of the rise in case counts and hospitalizations that started in November 2021, although as of early December 2021, observed trends largely were consistent with the uncertainty bounds.

State-level ensemble projections of cumulative cases for Scenario A (vaccination of children 5-11 years old without a new variant) over the period of September 12, 2021 to October 30, 2021 were well-correlated with reported cumulative cases (Pearson correlation coefficient *R* = 0.83, p<0.001) (Figure 2), although observed trends deviated from the model-projected decline in November 2021 (projection Pearson correlation coefficient 13 weeks into projection period *R* = 0.67, p<0.001). In particular, state-level observations tended to exceed ensemble projections for cumulative cases and deaths over this period. Based on ensemble projections, scenarios that assumed the emergence of a variant 50% more transmissible than the Delta variant in November 2021 led to models projecting a slow and moderate rise in national cases and deaths into early 2022 (Figure 1). In scenarios that did not assume the emergence of a new variant, models projected that national cases would drop to a level similar to that observed in June 2021, and projected deaths would drop to a weekly incidence of less than one per 100,000 individuals by March 12, 2022.

**Figure 2:**
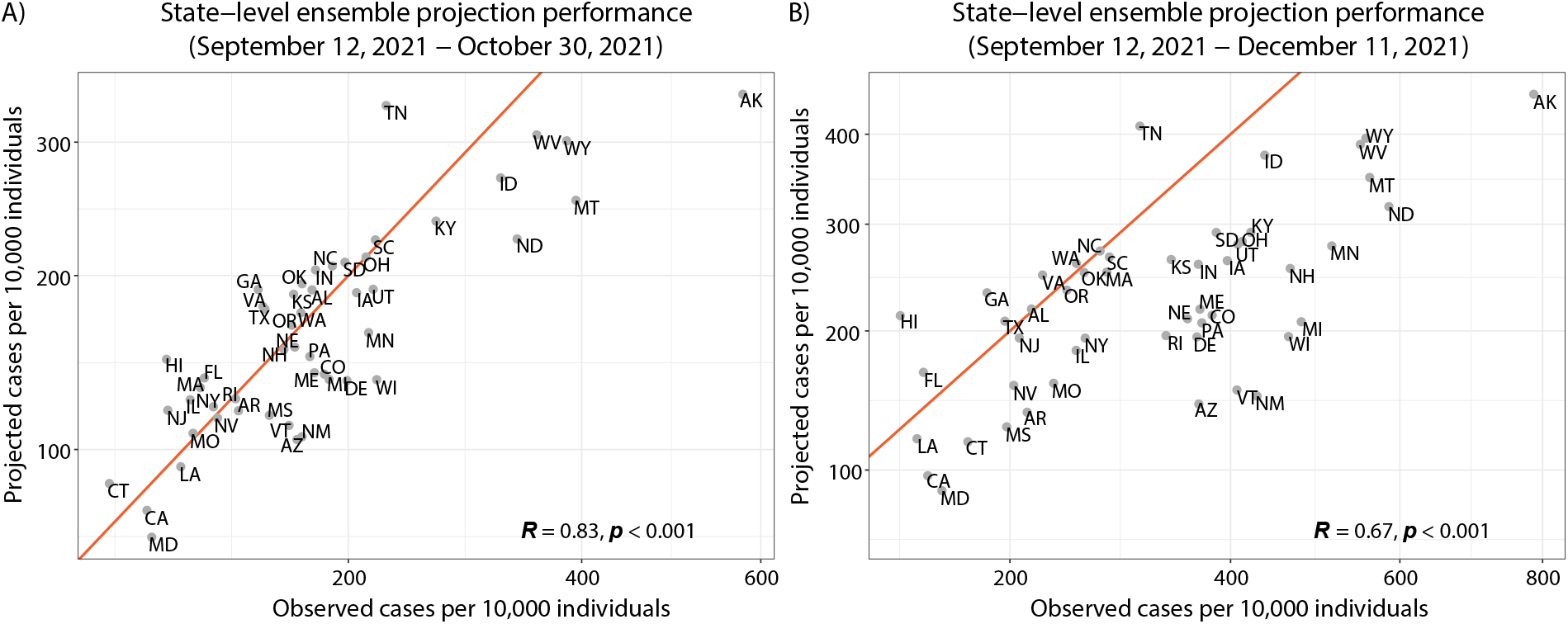
State-level ensemble projection performance from projection start date (September 12, 2021) through the observed weekly data available: A) before the assumed start of vaccination in 5-11 years old children (ending October 30, 2021; 6-week projection horizon) and after three months (ending December 11, 2021; 13-week horizon). Scenario A (childhood vaccination and no variant) projections are displayed. Projected cumulative cases vs. observed cumulative cases for all ages by state and normalized by state population. The red line marks where projected cumulative cases are equal to the corresponding state observations.

### Vaccine benefits

Assuming that vaccines showed strong effectiveness against the Delta variant and a hypothetical new variant, models projected that childhood vaccination would continue to reduce transmission burden across many important indicators (e.g., cases and hospitalizations). For the period November 1, 2021 to March 12, 2022, in the absence of a new variant, we estimated that vaccination of children 5-11 years old would avert ∼430,000 cases in the overall U.S. population (Figure 3), a 7.2% reduction (mean incidence ratio [IR] 0.928 95% confidence interval [CI] 0.880-0.977, Figure 4). With a more transmissible variant emerging in November 2021, the overall benefit of childhood vaccination increased to ∼860,000 cases averted, a reduction of 10.1% (mean IR 0.899, 95% CI 0.849-0.950). The models also projected that expanding vaccines to children could reduce hospitalizations by 11.8% (mean IR 0.882, 95% CI 0.805-0.959) with the introduction of a new variant and by 8.7% (mean IR 0.913, 95% CI 0.834-0.992) without, corresponding to absolute reductions of ∼93,000 and ∼47,000 hospitalizations, respectively. Similarly, vaccination of children 5-11 years old was projected to reduce overall population-level deaths by 12.3% (mean IR 0.877, 95% CI 0.759-0.994) in the presence of a new variant and by 9.2% (mean IR 0.908, 95% CI 0.797-1.020) without a new variant (Figure 3 and 4).

**Figure 3.**
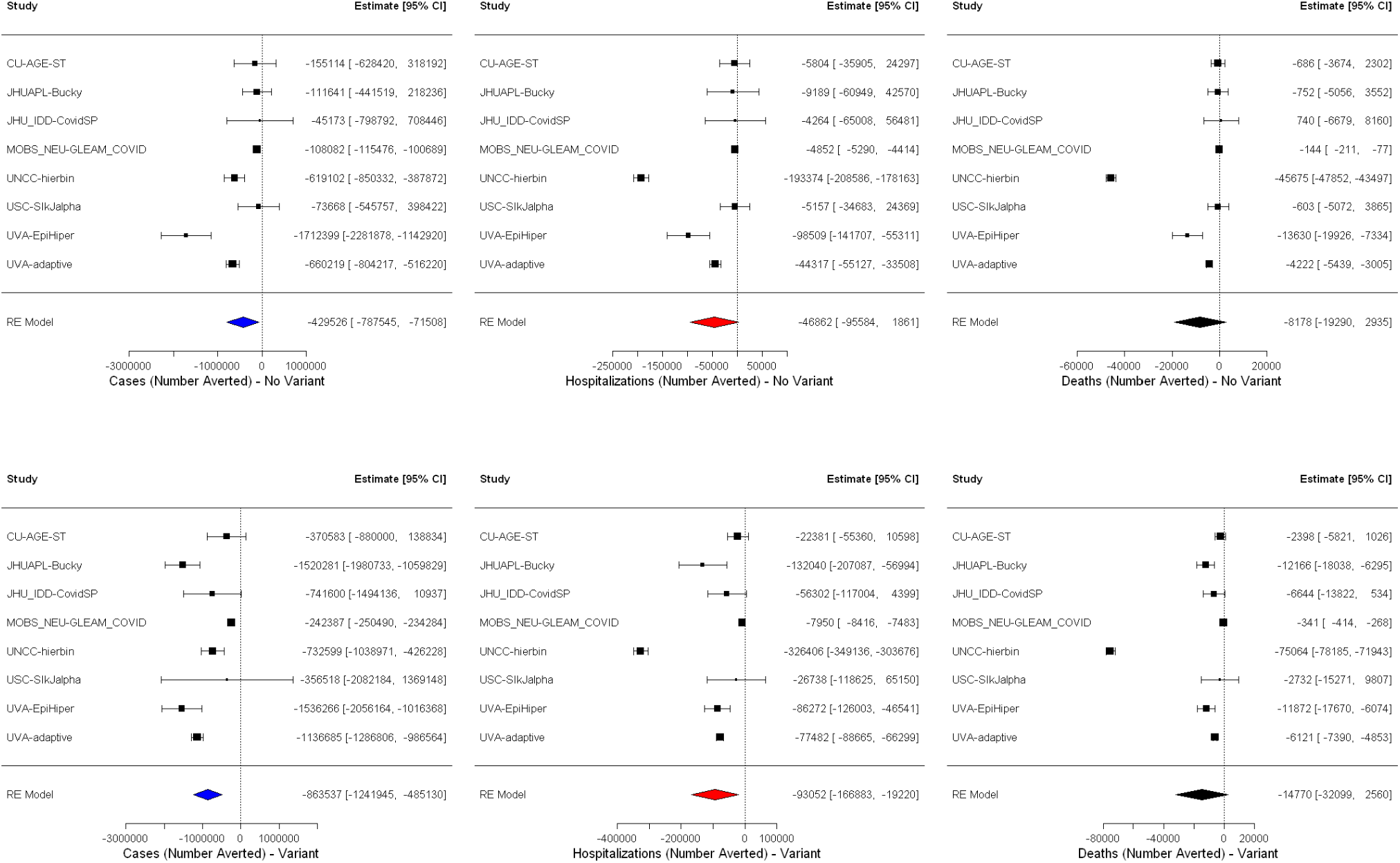
Scenario comparison. Absolute difference in estimates for all age cases, hospitalizations, and deaths when vaccination of 5-11-year-olds occurs without (top) and with (bottom) the emergence of a more transmissible variant from meta-analysis with random effects. Projection results from each team are analyzed as separate studies and are identified by team name abbreviation (see Supplemental Information for full team names).

**Figure 4.**
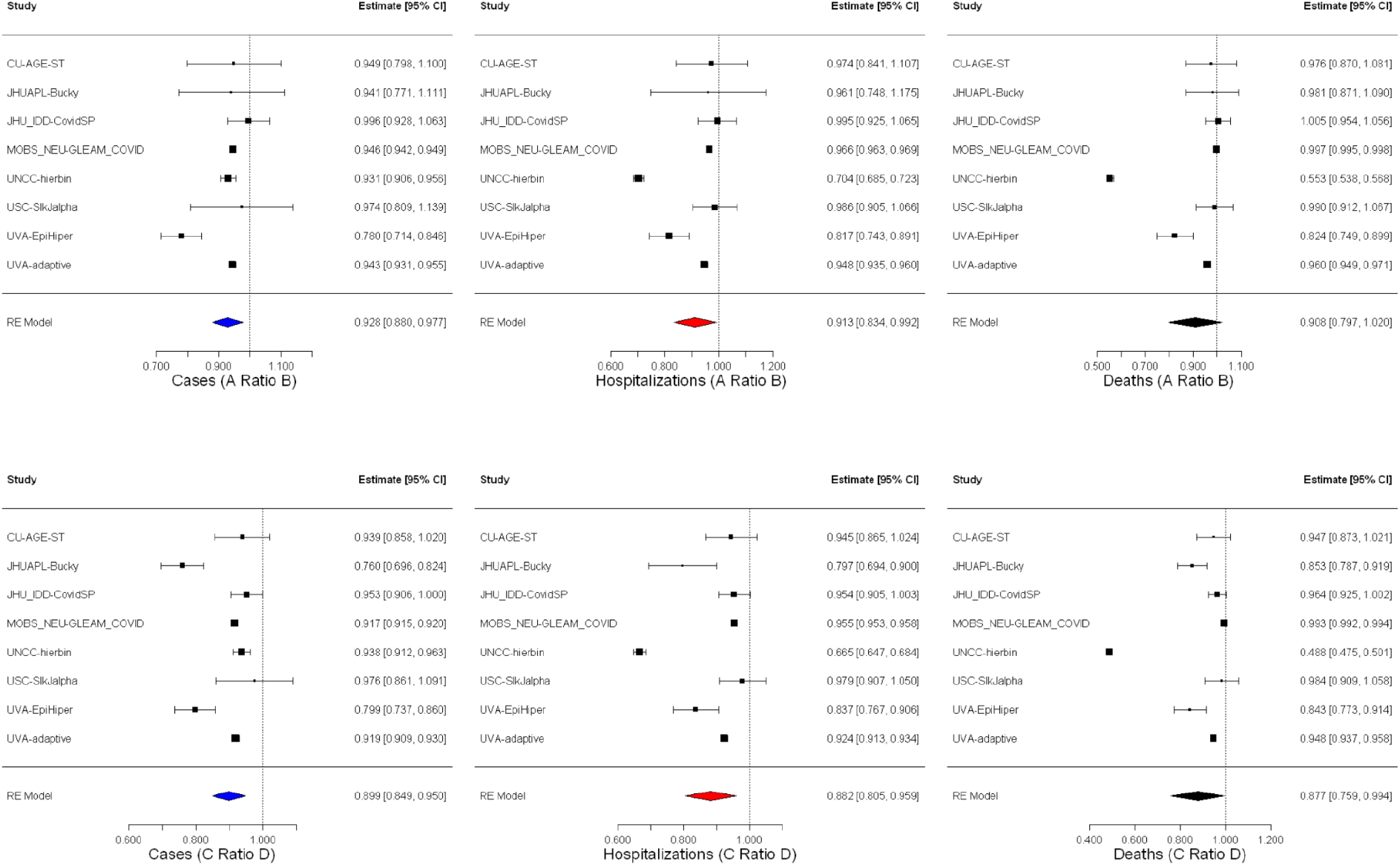
Scenario comparison. Incidence ratio estimates for all age cases, hospitalizations, and deaths when vaccination of 5-11year-olds occurs without (top – scenarios A and B) and with (bottom – scenarios C and D) the emergence of a more transmissible variant from meta- analysis with random effects. See Table 1 for scenario definitions.

At the state-level, the estimated effect of 5-11 year old vaccination varied substantially, with a median estimated reduction in reported cases of 5.8% (median IR 0.942, IQR 0.918-0.950) in the scenarios without the emergence of a more transmissible variant (range of reductions across states: -7.5-22.0%). Projected case reductions were more apparent at the state-level with the emergence of a highly transmissible variant, with a median reduction of 11.0% (reduction range -4.4-40.1%; median IR 0.890, IQR 0.818-0.914). Higher case reductions were projected to occur in states where vaccinated children 5-11 years old represent a higher proportion of the state population (Supplemental Figure S2).

Each of the five teams reporting younger age-group results projected that relative reductions in reported cases would be larger for the younger age group than overall (all age group) (Figure 5).

**Figure 5.**
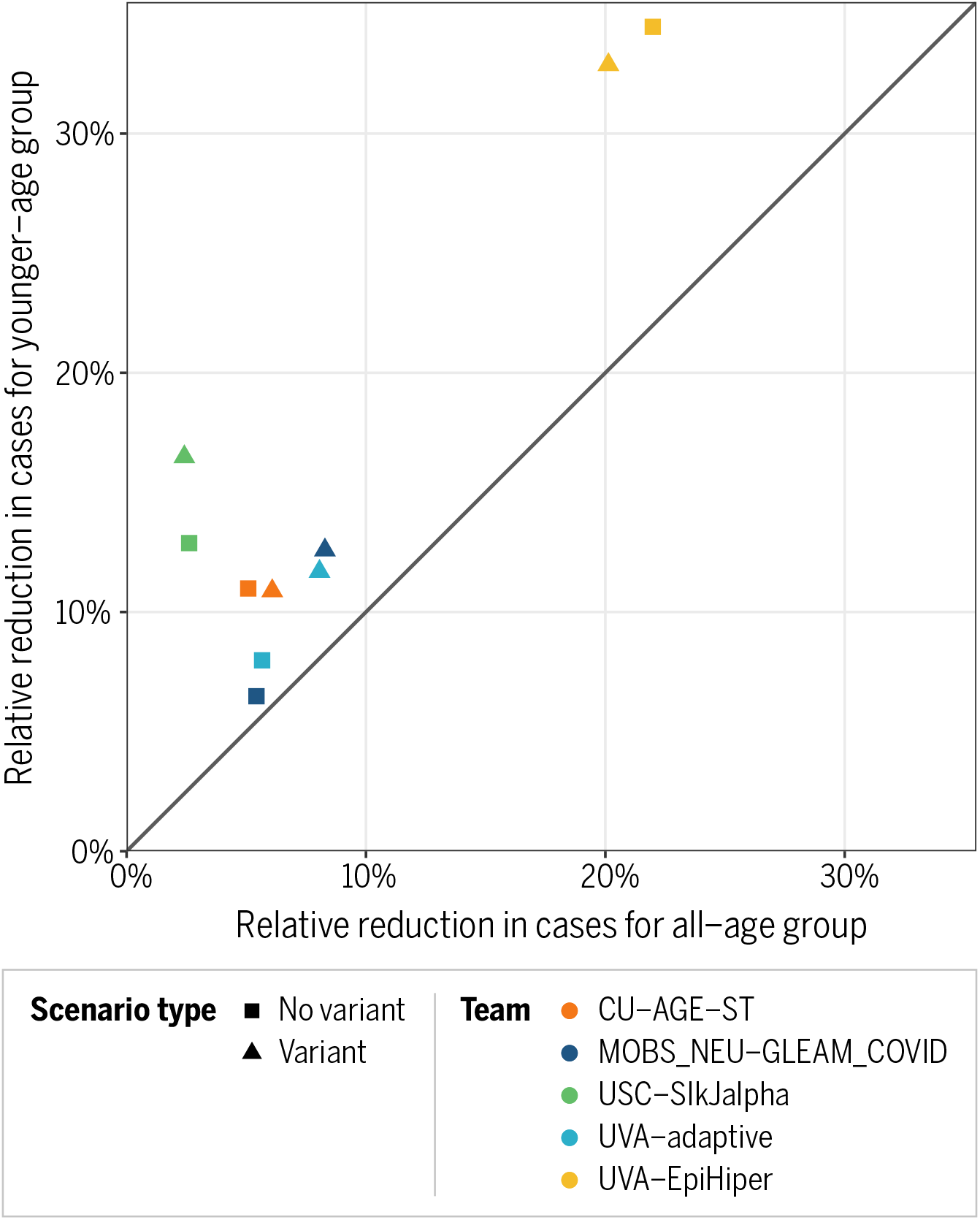
Comparison between younger-age projections and all-age projections for relative changes in cumulative cases where vaccination of 5-11-year-olds does not occur without (squares) and with (triangles) the emergence of a highly transmissible variant between November 1, 2021 and March 12, 2022. Younger-age groups considered are as follows: CU- AGE-ST (5-17 years), MOBS_NEU-GLEAM_COVID (0-11 years), USC-SlkJalpha (5-11 years), UVA-adaptive (0-17 years), UVA-EpiHiper (0-11 years).

## Discussion

Immunization of children 5-11 years old likely provides both direct and indirect benefits and provides protection against potentially more transmissible SARS-CoV-2 variants. Our multi-model effort projected continued declines in cases, hospitalizations and deaths through March 2022, under the assumption that a highly transmissible new variant did not emerge, among other assumptions. Under the scenarios where a hypothetical variant was assumed to emerge in November 2021, immunization of children 5-11 years old resulted in larger relative reductions in cases, hospitalizations and deaths among children than for the entire U.S. population, consistent with the existence of larger direct benefits to the age group vaccinated than (largely indirect benefits) overall.

Though immunizing 5-11 year-olds was projected by models to reduce cases by less than 10% overall (i.e., among all age groups), modest relative reductions translate to hundreds of thousands of cases, tens of thousands of hospitalizations, and thousands of deaths being averted. Population-level benefits are important because even though there is good direct protection afforded by the vaccine in most individuals, some groups (e.g., immunocompromised individuals, younger children, infants) rely on indirect protection^24^.

The new variant scenarios should be considered as a stress test against increases in SARS-CoV-2 transmissibility. Transmission can increase for a variety of reasons. Here we investigated a hypothetical variant with 50% increased transmissibility over the Delta variant and without immune escape (or changes in severity). Uncertainty remains surrounding Omicron, a variant which rapidly emerged in South Africa in mid-November 2021^25,26^ [Pulliam et al. medRxiv, Viana et al. Nature]. Although the Omicron variant likely has some degree of immune escape^25,26^ [Pulliam et al. medRxiv, Viana et al. Nature], increased intrinsic transmissibility may also contribute to its currently rapid spread^27^ [Yang and Shaman medRxiv 2021]. Prior work suggests that a moderate level of immune escape does not have substantial consequences unless paired with enhanced transmissibility, lending support to some transmission advantage^28^ [Bushman et al. Cell]. Further, differences in immunity acquired from natural infection or vaccination (and additionally by the type of vaccines and time since administration), the level of nonpharmaceutical interventions (NPIs), the prevalence of the Delta variant, and the severity of reinfections in different age groups, will affect the trajectory and burden of the Omicron variant in different populations. Despite differences between Omicron and our hypothetical variant, the result that vaccinating children 5-11 years old provides population-level benefits in terms of reductions in cases, hospitalizations and deaths, will likely hold. If the VE for symptomatic cases among children 5-11 years old was substantially lower than assumed by individual models in this set of projections, the population level and direct benefits realized would likely be lower.

This work was undertaken to both inform policy concerning approval and recommendation of vaccines in 5-11 year-olds and to complement communication efforts. At the population level, vaccine benefits are determined by vaccine coverage and effectiveness, modulated by the susceptibility of and transmissibility from the 5-11 years old age group. To provide timely projections (e.g., for public release on September 22, 2021 at https://covid19scenariomodelinghub.org/^12^<u>), several assumptions were made.</u>

Without known vaccine effectiveness in children at the time of projections, we assumed that vaccine effectiveness would be similar to that of adults, which may be somewhat conservative. Assumptions for VE against symptomatic Delta infection differed between models, ranging between 60-95% after two doses of mRNA vaccines. Recent data indicate a VE of 91% against symptomatic COVID-19 in 5-11 year olds^29^<u>[Pfizer briefing doc</u>], although no variant-specific VE estimate for 5-11 year-olds was available at the time of writing. Further, we assumed that immunization of children 5-11 years old would start on November 1, 2021; in fact the vaccine was approved on November 2 and some states expanded vaccine eligibility to 5-11 year-olds immediately following the approval. As regards vaccine coverage, we anticipated that more than 50% of U.S. children 5-11 years old would be vaccinated at the end of the projection period, guided by the uptake reported in adolescents during May-September 2021.

Our multi-model approach facilitates consideration of sources of epidemiological and situational uncertainty that affect projections of the direct and population-wide impacts of vaccination. The nine participating models made different assumptions regarding seasonality in transmission due to environmental conditions and behavior, waning of immunity, the expected levels of incidence in the coming months, and the potential role of children in transmission. A long-standing debate about age differences in susceptibility and transmissibility to COVID-19 persists, although there is ample evidence that children get infected and transmit SARS-CoV-2, as shown by numerous school outbreaks^30,31^ [Flasche S, Edmunds WJ. 2021, Torres et al. 2021]. Serology studies indicate that 38% of children 5-11 years old had been infected by SARS-CoV-2 prior to November 2021, confirming that infection frequently occurs in this age group^32^ [Jones 2021]. Overall, there was agreement between models in projecting higher vaccine benefits in periods of high incidence, as illustrated by the new variant scenarios. Accordingly, childhood vaccination is expected to help mitigate population-level surges in COVID-19 transmission.

Our study has several limitations. Only five modeling groups were able to provide age-resolved projections and the exact age groups differed between models due to different model structures and data inputs. Additionally, our results only considered health outcomes (cases, hospitalizations, and deaths) and did not consider other important outcomes such as missed days of work or school, or the cost of medical care. Accounting for these considerations would tend to increase the benefits of vaccination.

Additionally, we projected the benefits of vaccinating 5-11 year-olds on a relatively short time scale of a few months. Longer-term benefits are difficult to estimate as they depend on the balance between duration of immunity and viral evolution in different age groups. As we move into the next stages of the pandemic, with immunity increasing in all age groups through natural infection and vaccination, a shift to endemic dynamics with annual wintertime outbreaks is expected^33^<u>[Antia-Halloran]</u>. Incidence would also be projected to shift towards younger and immunologically naive individuals^34,35^ [<u>Lavine-Bjornstad-Antia,</u>, <u>Li-Metcalf-Bjornstad-Antia</u>], as is observed across a range of pathogens [e.g., Bansal et al.]. Based on observations of other pathogens^36,37^ [e.g., <u>Olson et al., Arinaminpathy et al.</u>], as the age distribution of disease becomes more concentrated in children, we expect direct and indirect vaccine benefits to increase in this age group.

Finally, the results in this paper are derived from scenario projections from a diverse set of individual models that are synthesized into a single projection. The accuracy of the conclusions rests on the validity of the counterfactual statements made by the ensemble projection. However, it is challenging to explicitly retrospectively evaluate scenario projections because the hypothesized scenarios will never come to realization exactly as assumed. For example, while our analyses suggest the true impacts might be higher in the presence of a new more transmissible variant, the characteristics of the emergent Omicron variant vary from the specifics of our scenarios, and thus should be interpreted with caution.

Increases in COVID-19 cases in the U.S. in January 2022 and worldwide indicate that the pandemic is not ending. This collaborative modeling effort underscored the large direct and overall benefits of expanding the vaccination program to 5-11 year-olds across a range of different scenarios, even including ones that produce gradual declines in COVID-19 incidences throughout mid-March 2022. Most importantly, increasing vaccination in children also builds resilience to potential increases in the upcoming pandemic trajectory, which may be fueled by new variants (e.g., Omicron), waning immunity, increased contacts, or other unpredictable factors.

## Data Availability

All projection results are publicly available. Other data referred to in the manuscript are from publicly available sources cited in the manuscript.

https://covid19scenariomodelinghub.org/

https://github.com/midas-network/covid19-scenario-modeling-hub

## Disclaimers

Any use of trade, firm, or product names is for descriptive purposes only and does not imply endorsement by the U.S. Government. The findings and conclusions in this report are those of the authors and do not necessarily represent the views of the National Institutes of General Medical Sciences, the National Institutes of Health, or the Centers for Disease Control and Prevention.

## Acknowledgments

K. Shea acknowledges support from NSF COVID-19 RAPID awards 2028301 and 2126278.

R. K. Borchering was funded by NSF COVID-19 RAPID award 2028301.

E. A. Howerton and K. Shea acknowledge support from the Huck Institutes for the Life Sciences at The Pennsylvania State University.

N. G. Reich was supported by the US CDC (1U01IP001122) and by the National Institutes of General Medical Sciences (R35GM119582).

L. Contamin, J. Levander, and J. Kerr, J. Espino, and H. Hochheiser were supported by NIGMS 5U24GM132013-02.

M. Kinsey, K. Tallaksen, S. Wilson, L. Shin, L. Mullany, K. Rainwater-Lovett were supported by HHS/ASPR Contract # 75A50121C00003.

M. Chinazzi, K. Mu, and A. Vespignani were supported by HHS/CDC 5U01IP0001137. J. Davis,

A. Pastore y Piontti, and A. Vespignani were supported by HHS/CDC 6U01IP001137.

A. Srivastava was supported by NSF, Grant No. 2027007.

S. A. Truelove, E. C. Lee, J. Lemaitre, C. Smith, J. Kaminsky, and J. Lessler acknowledge support from the Johns Hopkins Health System, the US Department of Health and Human Services / US Department of Homeland Security (DHHS/DHS), the State of California, the Johns Hopkins University Modeling and Policy Hub, and the Office of the Dean at the Johns Hopkins Bloomberg School of Public Health.

S. A. Truelove, E. C. Lee, J. Kaminsky, J. Dent, and C. Smith acknowledge support from NSF, Grant No. 2127976.

J. Lessler acknowledges support NIH Grant R01GM140564.

J. Lemaitre acknowledges support from the Swiss National Science Foundation.

J. Dent was supported by the State of California.

P. Porebski , S. Venkatramanan, A. Adiga, B. Lewis, B. Klahn, J. Outten, B. Hurt, H. Mortveit, A. Wilson, M. Marathe, J. Chen, S. Hoops, P. Bhattacharya, D. Machi acknowledge support from CDC grants 75D30119C05935 and U01CK000589, NSF grants CCF-1918656 and IIS-1931628, NIH grant 2R01GM109718-0, Dept of Defense – DTRA grant HDTRA1-19-D-0007, Virginia Dept Health awards VDH-21-501-0135 and VDH-21-501-0135, Virginia Dept of Emergency Management, UVA (internal seed grants). This research is based on survey results from Carnegie Mellon University’s Delphi Group.

G. España and A. Perkins received funding from an NSF RAPID grant (DEB 2027718).

S. Chen received funding from the Models of Infectious Disease Agent Study (MIDAS) Network (MIDASUP-05) and the North Carolina Biotechnology Center (2020FLG3898).

J. Shaman, S. Pei, T. Yamana and M. Galanti were supported by NIH Grant R01AI163023, CSTE Grant NU38OT000297 and a gift from the Morris-Singer Foundation. JS and Columbia University disclose partial ownership of SK Analytics. JS discloses consulting for Business Network International.

## Supplemental Methods

### COVID-19 Scenario Modeling Hub Round 9 Participating Teams

- Columbia University (New York, NY) – Age-Stratified Model (CU-AGE-ST)
- Johns Hopkins University (Baltimore, MD) ID Dynamics COVID-19 Working Group (JHU_IDD-CovidSP)
- Johns Hopkins University Applied Physics Lab (Laurel, MD) - Bucky (JHUAPL-Bucky)
- Northeastern University (Boston, MA) MOBS Lab – GLEAM COVID (MOBS_NEU- GLEAM-COVID)
- University of North Carolina at Charlotte (Charlotte, NC) – hierbin (UNCC-hierbin)
- University of Notre Dame (Notre Dame, IN) – FRED (NotreDame-FRED)
- University of Southern California (Los Angeles, CA) - SlkJalpha (USC-SlkJalpha)
- University of Virginia (Charlottesville, VA) – adaptive (UVA-adaptive)
- University of Virginia (Charlottesville, VA) – EpiHiper (UVA-EpiHiper)

### Projections

Probabilistic projections were collected using 23 quantiles (i.e., 0.01, 0.025, 0.05, every 5% to 0.95, 0.975, and 0.99).

Five teams did not include waning (CU-AGE-ST, JHUAPL-Bucky, JHU_IDD-CovidSP, UNCC- hierbin, NotreDame-FRED, see above list of participating teams for full team names). Three teams assumed that immunity would wane exponentially with an average period of either six months (UVA-EpiHiper) or one year (MOBS_NEU-GLEAM-COVID, UVA-adaptive). The USC- SlkJalpha team assumed a weighted average of assumptions about waning.

Five teams also submitted projections with the same quantile structure for a younger age-group (0-11, 5-11, 5-17, and 0-17 year groups) and an older age-group when possible:

- CU-AGE-ST
- MOBS_NEU-GLEAM_COVID
- USC-SlkJalpha
- UVA-adaptive
- UVA-EpiHiper

For scenarios without the emergence of a more transmissible variant, mean reported case reductions in younger age-groups ranged from 6.5 to 34.5% across models over the November 1, 2021 to March 12, 2022 time period. Reductions in hospitalizations and deaths ranged from 5.6 to 34.5% and 5.2 to 34.5%, respectively. For scenarios with the emergence of a more transmissible variant, mean projected reductions in cases were 10.9 to 32.9%, with hospitalization and death reductions ranging from 9.0 to 32.9% and 5.9 to 33.1% respectively. Percent reductions were not estimated for one team, which reported zero deaths for the younger age-group over the November 1, 2021 to March 12, 2022 time period.

### Meta analyses

To summarize the projected benefits of the vaccine program expansion across all modeling teams, we used a standard meta-analytic approach with random effects^22^. We estimated the mean difference in cumulative incidence and the mean incidence ratio between scenarios with and without children 5-11 years old vaccinated, stratified by presence or absence of the new variant, for the portion of the projection period following the assumed start date of childhood vaccination (i.e., from November 1, 2021).

Specifically, for each scenario *s* = *A, B, C, D* , the modeling teams *m* = 1,2,3 … 9 directly provided us with the the mean and variance (over their individual model replicates) of each cumulative outcome *o* = *Cases, Hospitalizations, Deaths*, at the start of the vaccination period (*t*_0_) and the end of the of projection period (*t*_1_) at the national level. We then estimated the mean (*µ*) and variance (σ^2^) for each model, scenario, and cumulative incidences over the period of interest as *μ*_*mso*_ = *μ*_*mso*_(*t*_1_) − *μ*_*mso*_(*t*_0_) and 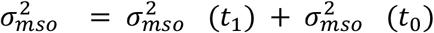 respectively. To compare scenarios, for example A (with vaccination of children 5-11 years old) and B (without vaccination of children 5-11 years old), we then estimated the mean of the difference as *µ*_*mso*_ − *µ*_*mBo*_ and the variance of the difference as 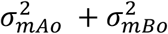. We also estimated the incidence ratio as the ratio of the above means (e.g. 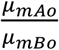, with the proportion reduction due to the vaccine estimated as 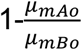); the variance of this ratio was obtained using the delta method. For both absolute difference and incidence ratio, we estimated the standard error (SE) as 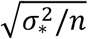, where 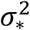 is the variance as defined above, and *n* is the number of replicates (simulations or sets of projected outcomes) for that model. Model specific means and standard errors were combined via random effects meta-analysis using restricted maximum likelihood (REML).

At the state level, individual modeling teams provided quantile distributions, as specified above, but did not provide us with model-specific estimates of mean and variance over their replicates at the two above time points of interest. Therefore, to evaluate vaccine benefits at the state level, we estimated the mean and variance from the 23 available points from each model-specific cumulative distribution function (CDF) at *t*_0_ and *t*_1_ (again, for each outcome, scenario, and state). Specifically, to each CDF (n∼11,000; 51 states x 9 models x 3 outcomes x 4 scenarios x 2 timepoints) we fit a penalized cubic spline Poisson regression model to estimate a continuous quantile function, from which we simulated 25,000 replicates. The mean and variance of these replicates were then estimated, and the above-described approaches were followed to obtain mean and variance for the absolute difference and incidence ratio between scenarios, and for combining these using random effects REML meta analyses. This procedure was also followed for approximating the direct effect of vaccine expansion (within the younger age group).

## Supplemental Figures/Tables

**Figure S1:**
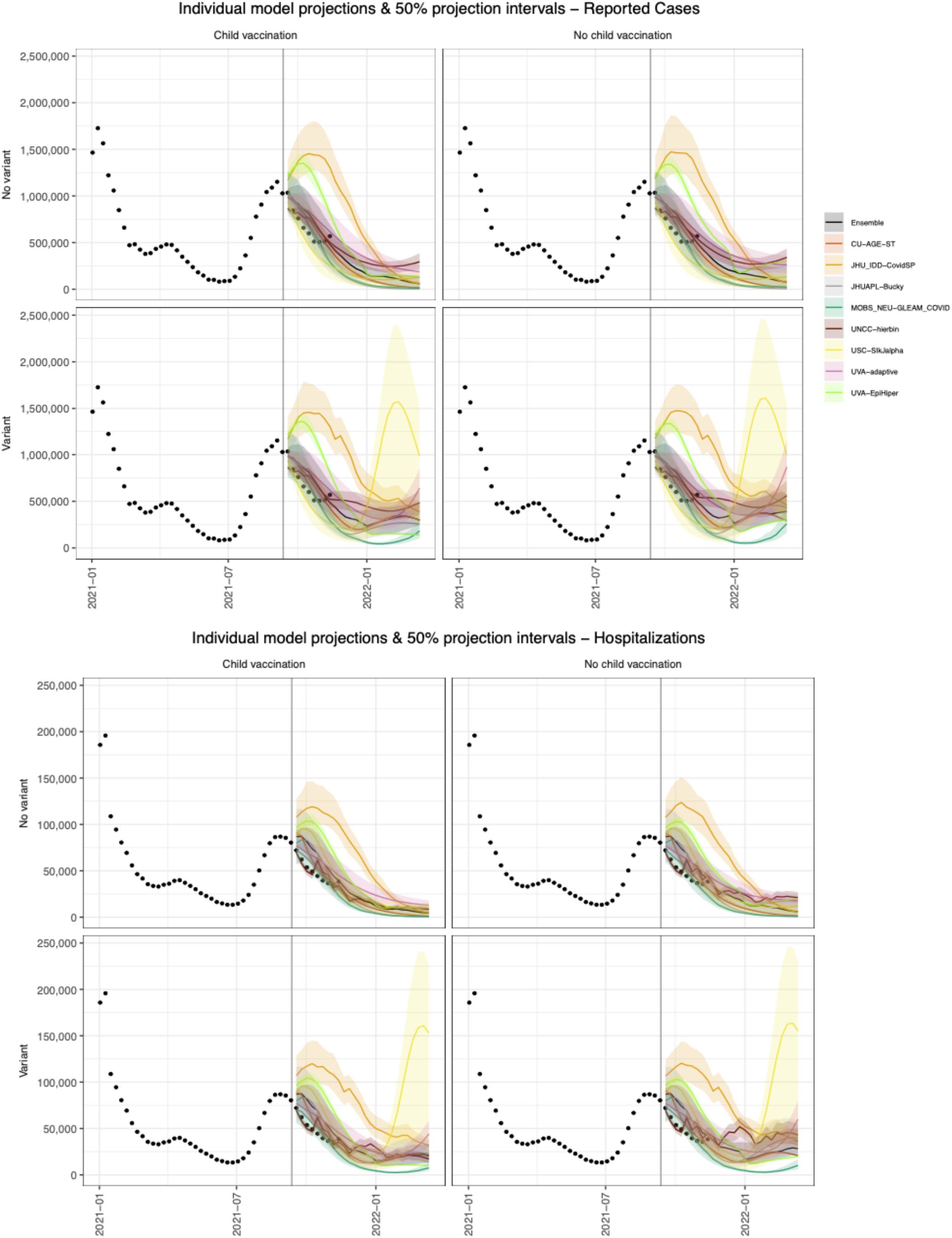
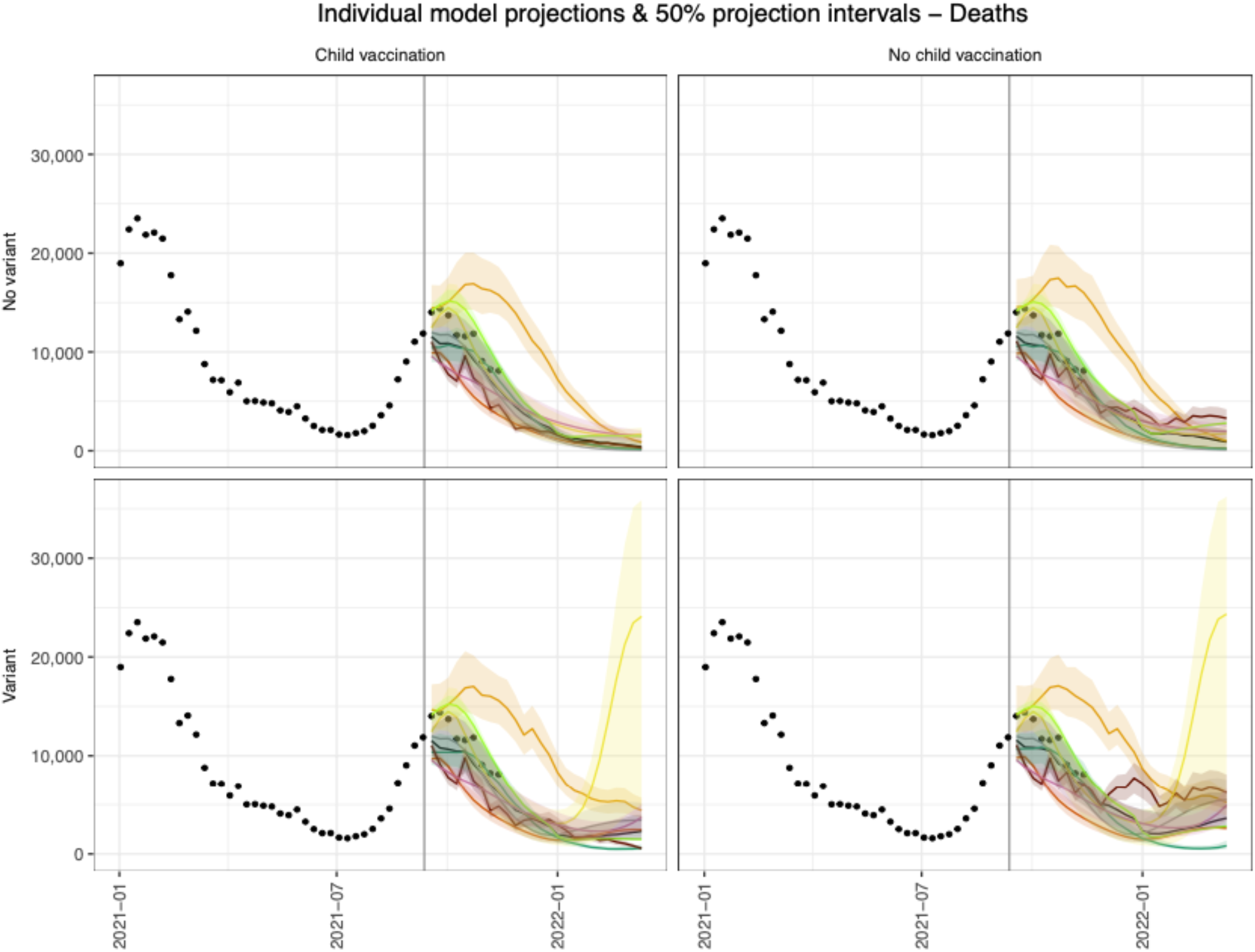
Model-specific trajectories for cases, hospitalizations, and deaths in each of the four scenarios. Differences between models were generally most apparent in the new variant scenarios. USC-SlkJalpha projected a large peak in cases in early 2022 in the variant scenarios. This peak was followed by peaks in hospitalizations and deaths. NEU-MOBs projected less of an impact in variant scenarios, with deep troughs observed at the national-level in early 2022. UVA-EpiHiper projected sharp decreases at the end of December 2021 and resurgences in the post-holiday period, driven by assumptions about school closures.

**Figure S2:**
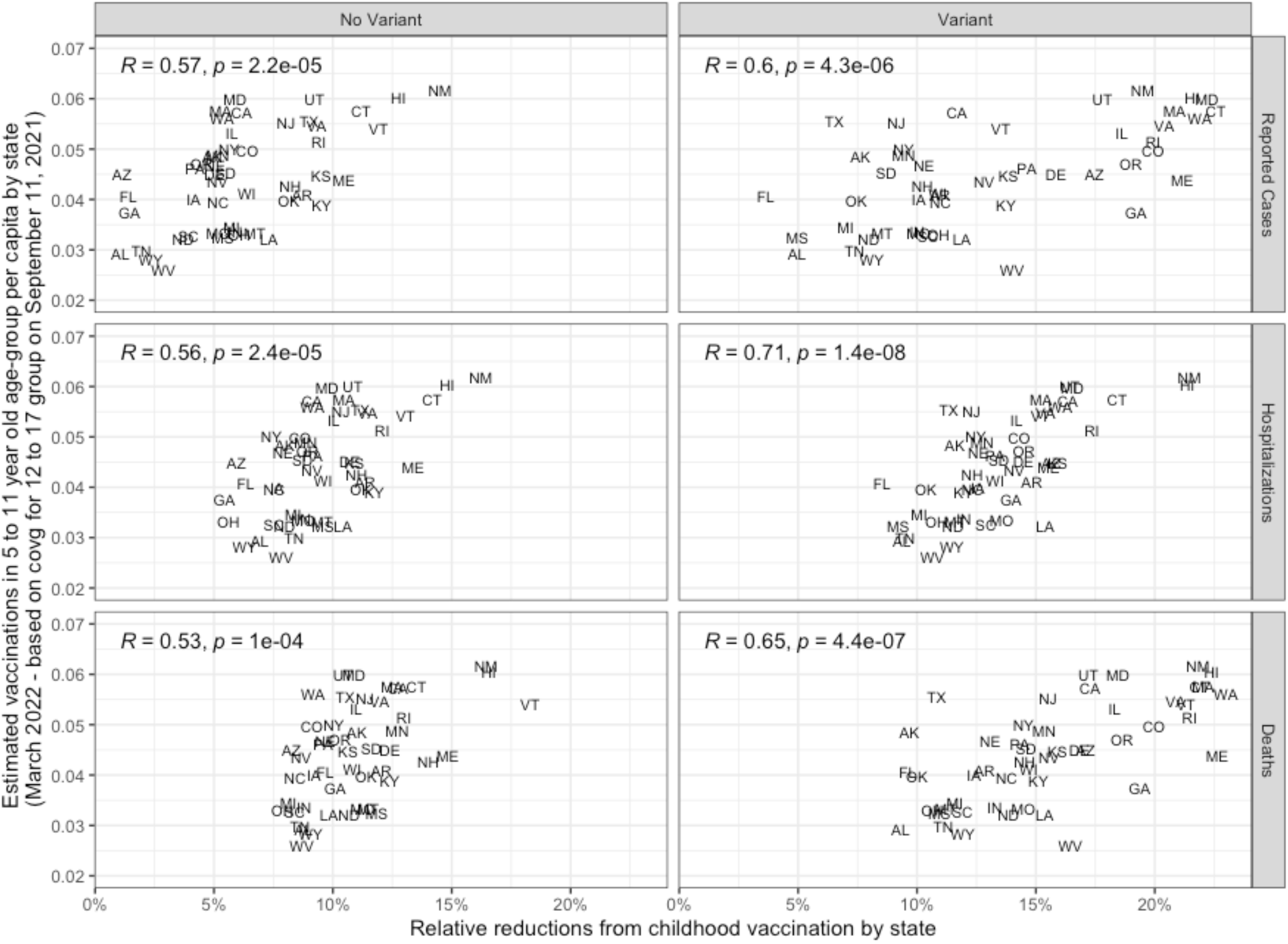
State-level estimates for number of vaccinated 5-11 year olds per capita vs. projected cases occurring between November 1, 2021 and March 12, 2022. Estimates for vaccinated 5-11 year olds are based on the vaccination coverage of 12-17 year olds on September 11, 2021 (the last day of data available for use in projections).

**Table S1:** model-specific vaccine effectiveness (VE) assumptions.

| Team | VE against infection | VE against symptoms | VE against hospitalization |
| --- | --- | --- | --- |
| CU-AGE-ST | 30% 2 wks after 1st dose,<br>70% 2 wks after 2nd dose | 30% 2 wks 1st dose,<br>70% 2 wks after 2nd dose | 90% 2 wks after 2nd dose |
| JHU-APL-Bucky | means of 35% after 1st dose,<br>85% after 2nd dose<br>(drawn from a distribution) | same as VE against infection | same as VE against infection |
| JHU_IDD-CovidSP | 35% 2wks after 1st dose,<br>60% 2wks after second | 35% 2wks after 1st dose,<br>60% 2wks after second | 80% 2wks after second dose |
| NotreDame-FRED | non-Delta: 76%, Delta: 72% | non-Delta: 90%, Delta: 85% | non-Delta: 96%, Delta: 96% |
| MOBS-NEU-GLEAM_COVID | 32% 2wks after 1st dose,<br>60% 2wks after 2nd dose | 35% 2wks after 1st dose,<br>85% 2wks after 2nd dose | same as VE against symptoms |
| UNCC-Hierbin | 60%-85% for all age groups and virus variants | same as VE against infection | 90% for all age groups and virus variants |
| USC-SlkJalpha | 85% minus waning | 85% minus waning | (100% minus waning) if in immune state |
| UVA-Adaptive | same as VE against symptoms | 50% after after 1st dose,<br>95% after second,<br>67% after J&J dose,<br>minus waning | same as VE against symptoms |
| UVA-EpiHiper | same as VE against symptoms | 35% after after 1st dose,<br>85% after second,<br>60% after J&J dose | same as VE against symptoms |

**Table S2:** additional model-specific assumptions including those about nonpharmaceutical interventions (NPIs).

|  | <u>CU-AGE-ST</u> | <u>JHUAPL Bucky</u> | <u>JHU ID D-CovidSP</u> | <u>NotreD ame-FRED</u> | <u>MOBS NEU - GLEAM CO VID</u> | <u>UNCC-hierbin</u> | <u>USC-SlkJalpha</u> | <u>UVA-adaptive</u> | <u>UVA-EpiHiper</u> |
| --- | --- | --- | --- | --- | --- | --- | --- | --- | --- |
| **Model Type** | Compartmental | Meta-population compartmental | Meta-population compartmental | Agent-based | Meta-population compartmental | Trajectory tracking (non-mechanistic) | Discrete time heterogeneous rate compartmental | Meta-population compartmental | Agent-based |
| **Geography** | County | County | State | State | County | State | State | County | State |
| **Mobility and contact data** | SafeGraph mobility, POLYMOD contact rates | SafeGraph mobility, age-based contact matrices | Commuting | Google mobility | Google mobility, commuting, flight, age-based contact matrices | Not used | Cuebiq contact scores data to model future NPI changes | Not used | ACS Commute, National Household Travel Survey |
| **Age groups** | 5 | 16 | Age-adjusted for vacc, hosp, and death | Individual ages | 10 | None | 10 | Hosp and death outcomes adjusted for 3 age groups | 5 |
| **Waning of natural immunity** | None | None | None | None | None | None | None | None | None |
| **Waning of vaccine immunity** | None | None | None | None | Leaky vaccine | None | Leaky vaccine | None | None |
| **Imports** | No | No | No | Low constant importations per variant | Domestic/International | None | No | Low steady importation per county, 1 per million population | None |
| **Multiple strains** | Proportion of variant cases by time imported from simplified two-strain model | Proportion of variant cases estimated from simplified model | No | Co-circulating variants matching dominance data. | 2 strains | No | Separate model for each strain | No | Two strains |
| **NPI history** | Fitted transmissibility; no additional NPI relaxation | Fitted based on transmission rates | Fitted based on state-level reported periods of changing policy | Fitted based on historic deaths | Mobility + Oxford data | Based on historical data | Cuebiq data to assess current contact scores | Modeled by proxy using fitted transmissibility | Not modeled |
| **NPI relaxation** | Maximum relaxation reached | Linear | Linear | NPIs relaxed for the projection period | Linear | NPIs held constant | Linear | Linear | NPIs held constant |
| **R0 wild strain** | Estimate in each county based on observed cases | 2.4 (mean) | 2.3 | Transmission-specific parameters adjusted to data. R0 was not specified explicitly | Prior in [1.6, 3.8]. Posterior defined by the model calibration in each state. | NA (due to non-mechanistic nature of the model) | Estimated from current data for current contact scores, and adjusted to pre-COVID contact scores | Variable, as we estimate intrinsic transmissibility and NPI effects through fitting of transmission rates from observed case series | NA (wild type not explicitly modeled, model initialized with delta variant with calibrated R_effective which varies among states) |

Note: additional assumptions that are left at the teams' discretion are unspecified here to encapsulate current sources of scientific uncertainty (e.g., transmissibility assumptions).

## Notes

### Competing Interest Statement

JL has served as an expert witness on cases where the likely length of the pandemic was of issue. MCR reports stock ownership in Becton Dickinson & Co., which manufactures medical equipment used in COVID-19 testing, vaccination, and treatment. JS and Columbia University disclose partial ownership of SK Analytics. JS discloses consulting for BNI. There are no other competing interests to declare.

### Funding Statement

K. Shea acknowledges support from two National Science Foundation (NSF) COVID-19 RAPID awards 2028301 and 2126278. R. K. Borchering was funded by NSF COVID-19 RAPID award 2028301. E. A. Howerton and K. Shea acknowledge support from the Huck Institutes for the Life Sciences at The Pennsylvania State University. N. G. Reich was supported by the US CDC (1U01IP001122) and by the National Institutes of General Medical Sciences (NIGMS) (R35GM119582). L. Contamin, J. Levander, and J. Kerr, J. Espino, and H. Hochheiser were supported by NIGMS 5U24GM132013-02. M. Kinsey, K. Tallaksen, S. Wilson, L. Shin, L. Mullany, K. Rainwater-Lovett were supported by the U.S. Department of Health and Human Services (HHS) Office of the Assistant Secretary for Preparedness and Response (ASPR) HHS/ASPR Contract # 75A50121C00003. M. Chinazzi, K. Mu, and A. Vespignani were supported by HHS/CDC 5U01IP0001137. J. Davis, A. Pastore y Piontti, and A. Vespignani were supported by HHS/CDC 6U01IP001137. A. Srivastava was supported by NSF Grant No. 2027007. S. A. Truelove, E. C. Lee, J. Lemaitre, C. Smith, J. Kaminsky, and J. Lessler acknowledge support from the Johns Hopkins Health System, the US Department of Health and Human Services / US Department of Homeland Security (DHHS/DHS), the State of California, the Johns Hopkins University Modeling and Policy Hub, and the Office of the Dean at the Johns Hopkins Bloomberg School of Public Health. S. A. Truelove, E. C. Lee, J. Kaminsky, J. Dent, and C. Smith acknowledge support from NSF, Grant No. 2127976. J. Lessler acknowledges support the National Institutes of Health (NIH) Grant R01GM140564. J. Lemaitre acknowledges support from the Swiss National Science Foundation. J. Dent was supported by the State of California. P. Porebski , S. Venkatramanan, A. Adiga, B. Lewis, B. Klahn, J. Outten, B. Hurt, H. Mortveit, A. Wilson, M. Marathe, J. Chen, S. Hoops, P. Bhattacharya, D. Machi acknowledge support from CDC grants 75D30119C05935 and U01CK000589, NSF grants CCF-1918656 and IIS-1931628, NIH grant 2R01GM109718-0, Dept of Defense - DTRA grant HDTRA1-19-D-0007, Virginia Dept Health awards VDH-21-501-0135 and VDH-21-501-0135, Virginia Dept of Emergency Management, UVA (internal seed grants). This research is based on survey results from Carnegie Mellon University Delphi Group. G. España and A. Perkins received funding from an NSF RAPID grant (DEB 2027718). S. Chen received funding from the Models of Infectious Disease Agent Study (MIDAS) Network (MIDASUP-05) and the North Carolina Biotechnology Center (2020FLG3898). J. Shaman, S. Pei, T. Yamana and M. Galanti were supported by NIH Grant R01AI163023, CSTE Grant NU38OT000297 and a gift from the Morris-Singer Foundation. JS and Columbia University disclose partial ownership of SK Analytics. JS discloses consulting for BNI.

### Author Declarations

This analysis was conducted using publicly available, aggregated data on cases, hospitalizations, and deaths, along with published parameter estimates. No human subjects research was involved, and no IRB approval was required.

